# Multifocal vascular affection in a patient with angina pectoris - rationale and design of a clinical study

**DOI:** 10.1101/2021.03.23.21253594

**Authors:** M. Pintova, I. Simova, N. Dimitrov, J. Krasnaliev, V. Kornovski, T. Vekov

## Abstract

Atherosclerosis is a disease of the large and middle arteries and is characterized mainly by endothelial dysfunction, inflammation of the vascular wall and the accumulation of lipids, cholesterol, calcium and cell debris in the intima of the vascular wall. This accumulation leads to plaque formation, vascular remodeling, acute and chronic obstruction of the vessel lumen, blood flow abnormalities, and reduced oxygen supply to target organs. As a result, blood vessels become harder and their lumen shrinks, which increases the risk of obstruction and thrombosis. Depending on the affected area, the corresponding symptoms appear - angina or myocardial infarction when affecting the coronary circulation; transient ischemic attack (TIA) or stroke affecting cerebral circulation; claudication or gangrene when affecting the limbs; aneurysm or dissection affecting the aorta. Various risk factors are known to affect the onset of the disease. They are divided into adjustable (dyslipoproteinemia, hypertension, diabetes mellitus, smoking, obesity, inactivity, emotional stress, etc.) and unadjustable (gender, age, family predisposition). The risk of developing atherosclerosis increases significantly in the presence of several risk factors. For example, the presence of 2 risk factors increases the risk of developing atherosclerosis 4 times.

The atherosclerotic process is multifocal - it develops everywhere in the body - affecting the cardiac, cerebral and peripheral arteries almost simultaneously or sequentially. Its isolated manifestations in only one area are an exception.

## Introduction

Atherosclerosis is often generalized. Depending on the degree of involvement of the various organs by atherosclerosis, its marks also appear. When atherosclerosis severely affects the blood vessels of the heart, symptoms of angina and heart attack appear. When the cerebral arteries are affected, cerebral atherosclerosis develops, which gives many and varied signs - disturbed memory (especially for recent events, while memories of the distant past are well preserved).

Patients become slower, move in small steps. Their character is changing. They complain of headache, dizziness, sleep disturbance. In severe cases, the cerebral thrombosis can develop with paralysis of a certain group of muscles, speech disorders, etc.

Atherosclerosis of the lower extremities disrupts their blood supply and leads to tingling, muscle weakness and pain. Typical are the pains when walking, which force the patient to stop and rest. In more severe cases, gangrene of the limb may develop. (1,2)

In June 2020 at Heart and Brain Center of Excellence, University Hospital – Pleven, Bulgaria we began monitoring patients referred for selective coronary arteriography, in order to establish accompanying atherosclerotic changes in other major arterial vessels.

## Rationale for the study

Patients with atherosclerotic involvement of an area are at increased risk of fatal and non-fatal cardiovascular (CV) events

When a vascular area is affected by atherosclerosis, it not only threatens the relevant organ (e.g. the brain in carotid arterial disease), but also increases the overall risk of any type of CV event (e.g. coronary events). Any vascular area affected by atherosclerosis can be considered a marker of realized CV risk.

Atherosclerosis manifests itself in several clinical forms, distinguishing coronary artery disease, aortic disease and peripheral arterial disease (cerebrovascular disease, upper extremity artery disease, visceral atherosclerosis, lower extremity artery disease).

**Figure 1.**
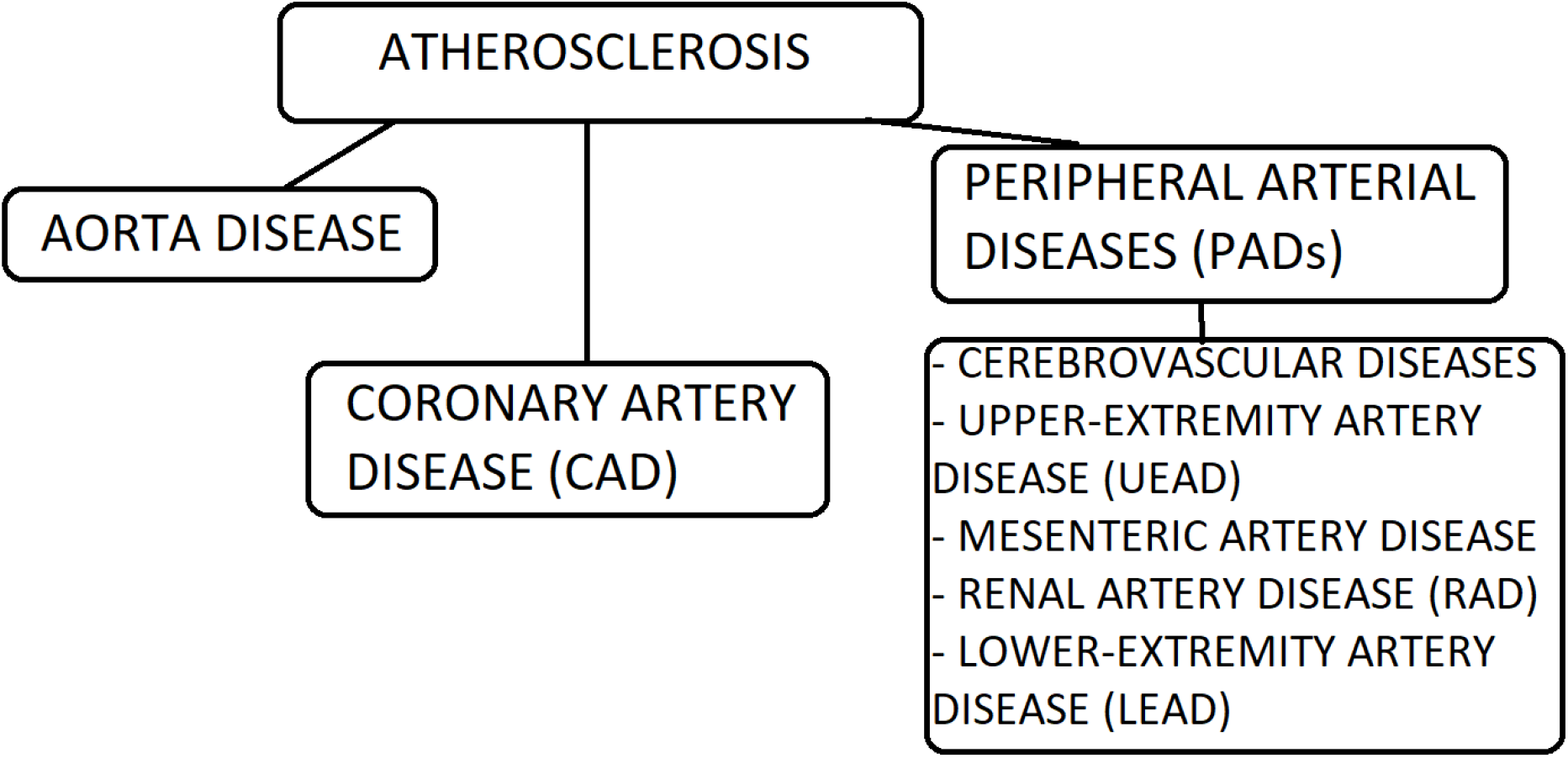
Clinical forms of atherosclerosis.

Ischemic heart disease caused by coronary atherosclerosis affects about 5% of the population, and at the age of 65 this frequency reaches 20%.

Cerebrovascular atherosclerosis affects the carotid, vertebral and intracranial arteries and causes ischemic stroke. In Bulgaria, the mortality rate from stroke is higher than that from myocardial infarction. Ischemic stroke causes severe damage to health. In its acute phase, one third of patients die, then one third remain disabled, and one fifth receive a stroke within a year.

Peripheral arterial disease of the limbs - PAD covers about 4% of the population, with 12 million people in the United States and 20 million in Europe. For Bulgaria, the probable number is 300,000 people. PAD affects the lower and upper limbs and causes varying degrees of disability, and in severe cases - gangrene. PAD is a deadly disease. By the 5th year, 30% of patients die from other forms of atherosclerosis - heart attack and stroke. Critical limb ischemia results in 30% amputations and 20% mortality. (12)

## Materials and methods

In our study we included patients admitted to the Cardiology Clinic for angina complaints and indicated for selective coronary arteriography. In them, we monitored the involvement of the carotid arteries by ultrasound (US), and the determination of the ankle brachial index (ABI) was the method of choice in determining the presence of atherosclerosis in the arteries of the lower extremities.

Selective coronary arteriography - the golden standard for establishing coronary artery disease. This is an invasive X-ray contrast study. It is usually performed as part of cardiac catheterization, which involves left ventricular angiography with measurement of hemodynamic parameters, providing a more complete and comprehensive examination of individual CV status. Invasive coronary angiography is routinely performed relatively safely.

The vascular access is arterial - a.femoralis, a.radialis or a.brachialis. Historically, the first method used was through a.femoralis. Femoral access is inappropriate in patients who are unable to be immobilized for a long period of time, such as those at high risk of bleeding or with peripheral arterial disease. Recently, radial access is most often used, providing greater comfort for the patient. Brachial access is appropriate in patients who have difficulty performing angiography due to vascular tortuosity, obesity, or in whom other vascular access is not possible.

Different catheters are used depending on their length, diameter and shape. The most commonly used catheters for diagnostic coronary angiography are Judkins and Amplatz catheters. The first is suitable for both femoral and radial access. The second type is a good alternative to Judkins catheters.

They are suitable for cannulation of the right coronary artery and sinus venosus grafts (SVGs). Multipurpose (MP) catheters are suitable for hard-to-reach coronary artery ostiums or SVGs.

In order to determine the extent of a coronary lesion, it is necessary to properly visualize the coronary circulation. There are several basic projections that are used. This is done by proper angulation of the X-ray tube of the angiograph.

**Table 1.** Angiographic projections of the coronary arteries.

| ***Left coronary artery*** | ***Anatomic description*** |
| --- | --- |
| AP - 5-10° RAO | LM |
| 30-45° LAO + 20-30° cranial | LAD-LCx bifurcation |
| 30-40° RAO + 20-30° caudal | LCx + OMs |
| 5-30° RAO + 20-45° cranial | LAD+Diag |
| 50-60° LAO + 10-20° caudal (spider) | LAD-LCx bifurcation, LCx, OMs |
| ***Right coronary artery*** | ***Anatomic description*** |
| 30-45° LAO + 15-20° cranial | Prox, Mid, PDA |
| 30-45° RAO | Prox, Mid, PDA |

LM – Left main; LAD – Left anterior descending; LCx – Left circumflex; OM – Obtuse marginal; PDA – Posterior descendent artery

**Figure 2.**
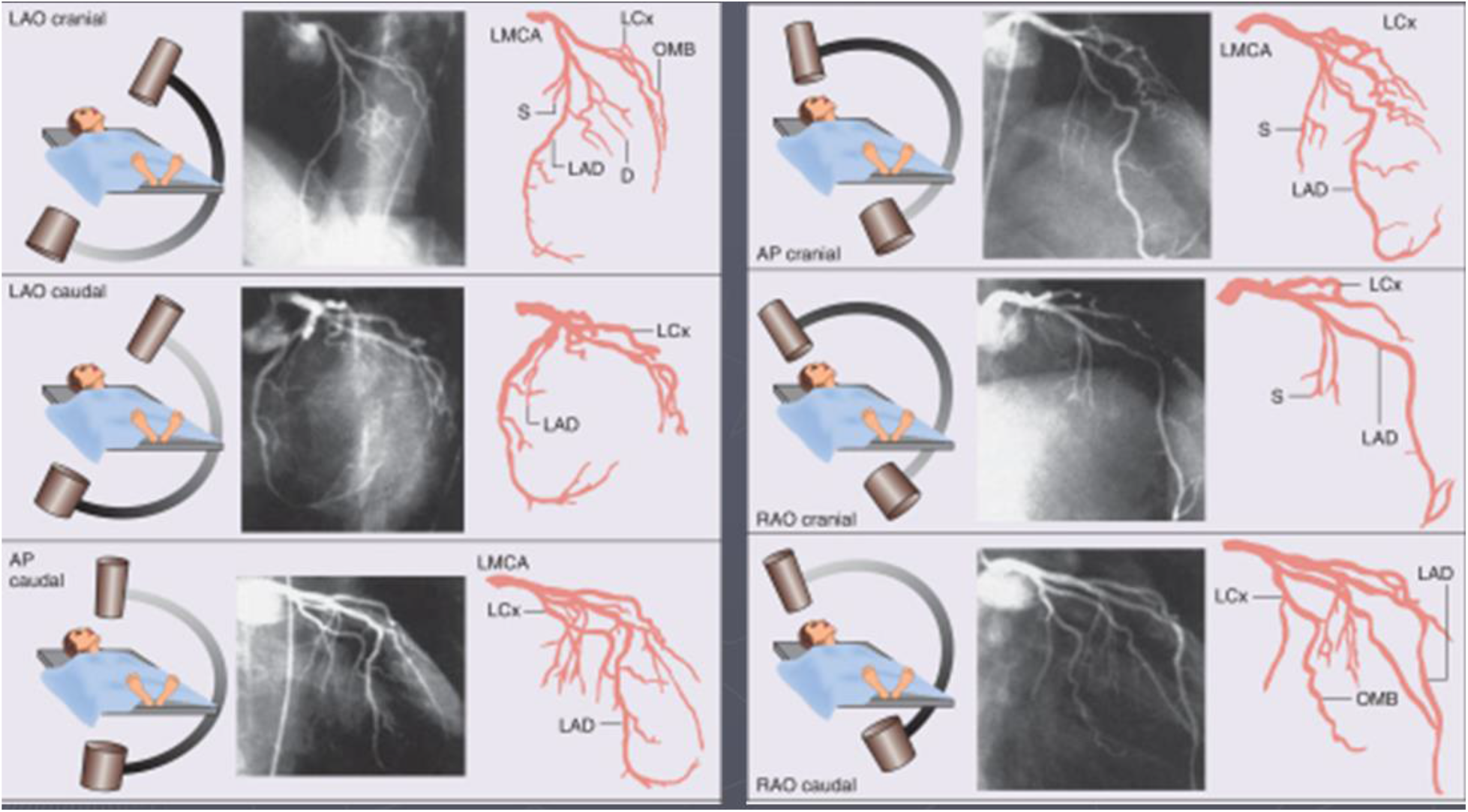
Angiographic projections for left coronary artery (LMCA)

**Figure 3.**
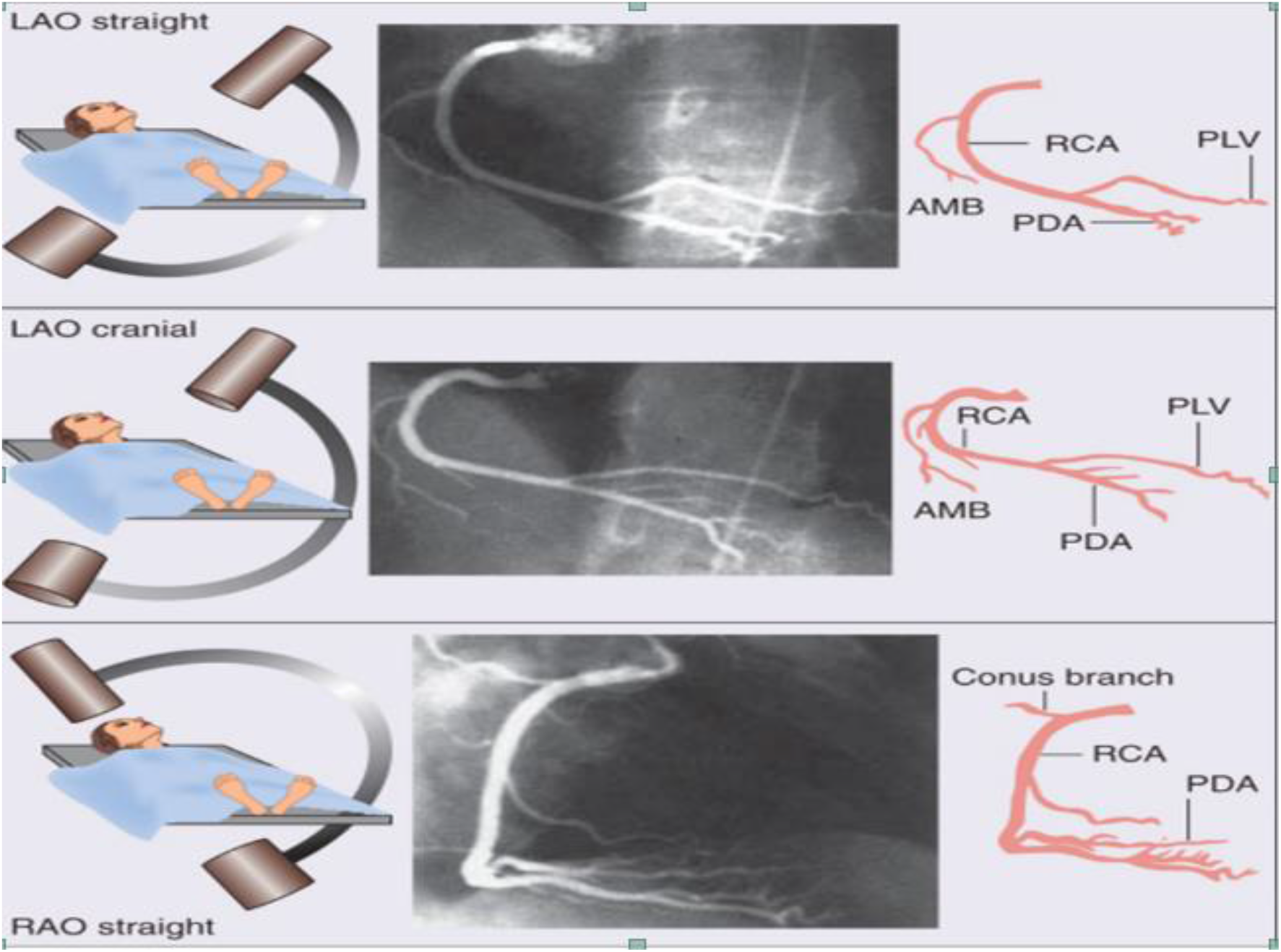
Angiographic projections for right coronary artery (RCA) (1, 7, 8, 13)

Ultrasound of carotid arteries - a highly effective method of diagnosis that allows to identify a range of structural changes in the carotid arteries: atherosclerotic lesions, stenosis (narrowing), vascular obstruction (including associated with osteochondrosis), abnormalities in the development of blood vessels and others.

Procedure in two stages (duplex): - Representation of the vessel: B-mode (brightness) technique; depiction of the vessel in black and white scale (grayscale image); color Doppler imaging can be superimposed on this image; - Spectral analysis: allows assessment of blood flow velocity; both visual and audible signal;

Visual determination of the degree of stenosis is not reliable enough; • The most commonly used methods for acoustically determining the degree of stenosis include: - Measurement of peak systolic (PSV) and telediastolic velocity (EDV); - Defining ratios, eg: Internal carotid artery (ICA) PSV / Common carotid artery (CCA) PSV

**Table 2.** Evaluation of ICA stenosis severity.

| Degree of stenosis (NASCET) | PSV – ICA (m/s) | Plaque estimate (%) | ICA/CCA PSV ratio | EDV- ICA (m/s) |
| --- | --- | --- | --- | --- |
| Normal | <125 | None | <2 | <40 |
| <50% | <125 | <50% | <2 | <40 |
| 50-69% | 125 - 230 | >50% | 2-4 | 40-100 |
| >70% | > 230 | >50% | >4 | >100 |
| Near occlusion | Variable | Visible | Variable | Variable |
| Total occlusion | Undetectable | No lumen | N/A | N/A |

Normal: ICA PSV < 50%
Stenosis: ICA PSV 70%
stenosis to subtotal occlusion: ICA PSV > 230 cm / s in the presence of visible plaque and narrowing of the lumen;
Subtotal occlusion: marked narrowing of the lumen in an image with color Doppler;
Complete occlusion: no lumen of the black and white scale is visualized and no blood flow of color Doppler, spectral Doppler and Power Doppler is detected

ABI - ankle-brachial index - noninvasive, reliable measurement for ishcaemia of the lower limbs. ABI is tested according to standard methods.

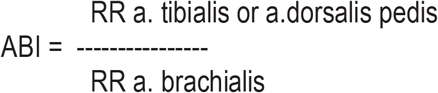

Where ABI is ankle-brachial index;

RR a. tibialis - the higher systolic pressure of one of the two step arteries;

RR a. brachialis - the higher systolic pressure of one of the two brachial arteries.

The normal value is 1.0 ± 0.1. There are variations in the value of segmental blood pressure, but ABI <0.90 is always abnormal. Such values most likely indicate peripheral arterial stenosis ≥50% (sensitivity - 90% and specificity - 98%).

ABI is a criterion for determining the severity of lower extremity artery disease (LEAD). Its average values for determining the different stages of LEAD are:

Normal - 1.09;

I-IIA stages - 0.8;

II B stages - 0.56;

Stages III and IV - 0.23.

(1,2,12)

### Study design

The study was initiated in June 2020 at Heart and Brain Center of Excellence, University Hospital – Pleven, Bulgaria. Patients hospitalized in the Cardiology Clinic were referred for invasive assessment of coronary circulation in view of manifested angina symptoms or equivalent. Patients who got involved were of different ages, men and women with different social status and different risk factors for atherosclerosis - dyslipidemia, diabetes, smoking, obesity, family history.

Cases of patients hospitalized on another occasion - exacerbated heart failure, arrhythmia and pulmonary embolism - were excluded from the present analysis.

All patients had a detailed history with physical status, and transthoracic echocardiography was performed to assess cardiac function. In parallel, ultrasonographic examination of the carotid arteries and measurement of the ABI index were performed. The patients were then referred for selective coronary arteriography.

### Ethical considerations

All patients sign an informed consent for selective coronary arteriography. All patients are explained the benefit of screening for atherosclerotic involvement in other vascular areas, for which they give their agreement. All patients sign an informed consent for the processing and storage of personal data. The study is in line with the Declaration of Helsinki.

### Expected results

We expect to follow more than 200 patients by the end of March 2021. The study will allow us to assess what part of patients with angina have concomitant cerebrovascular disease and / or PAD. In addition, we will analyze the incidence of cerebrovascular disease and / or PAD in patients with microvascular coronary heart disease, as well as those with one-, two- and three-clone coronary heart disease.

The described analysis will allow us to build an algorithm for screening patients with angina with a view to early detection and timely treatment of concomitant cerebrovascular disease and / or PAD.

## Discussion

Atherosclerosis is a systemic progressive process that affects simultaneously or sequentially different vascular regions, lasting asymptomatically or with discrete complaints for a long time, until the onset of a severe or fatal complication.

The widespread prevalence of combined cardiovascular and peripheral vascular disease has been confirmed in two major international studies - the REACH (Reduction in Atherothrombosis for Continued Health) registry and AGATHA (A Global Atherothrombosis Assessment), in which 16-35% of patients (with atherosclerosis or ≥ 3 risk factors) have polyvascular disease [3, 4]. Significant coronary stenosis in at least one vessel is found in about 60% of patients with severe peripheral vascular disease requiring surgery[5]. The presence of combined vascular pathology is associated with an almost doubling of mortality to 4.6% per year, compared with the single manifestation of cardiac or peripheral atherosclerosis [3]. The one-year risk of myocardial infarction, ischemic stroke, or hospitalization for atherothrombotic events among these patients was 23.1% (compared with 13–17% in single disease) according to the REACH registry [3]. Peripheral vascular disease is a more important risk factor for predicting mortality than previous myocardial infarction or the severity of angina, according to the CASS registry. (10)

**Figure 4.**
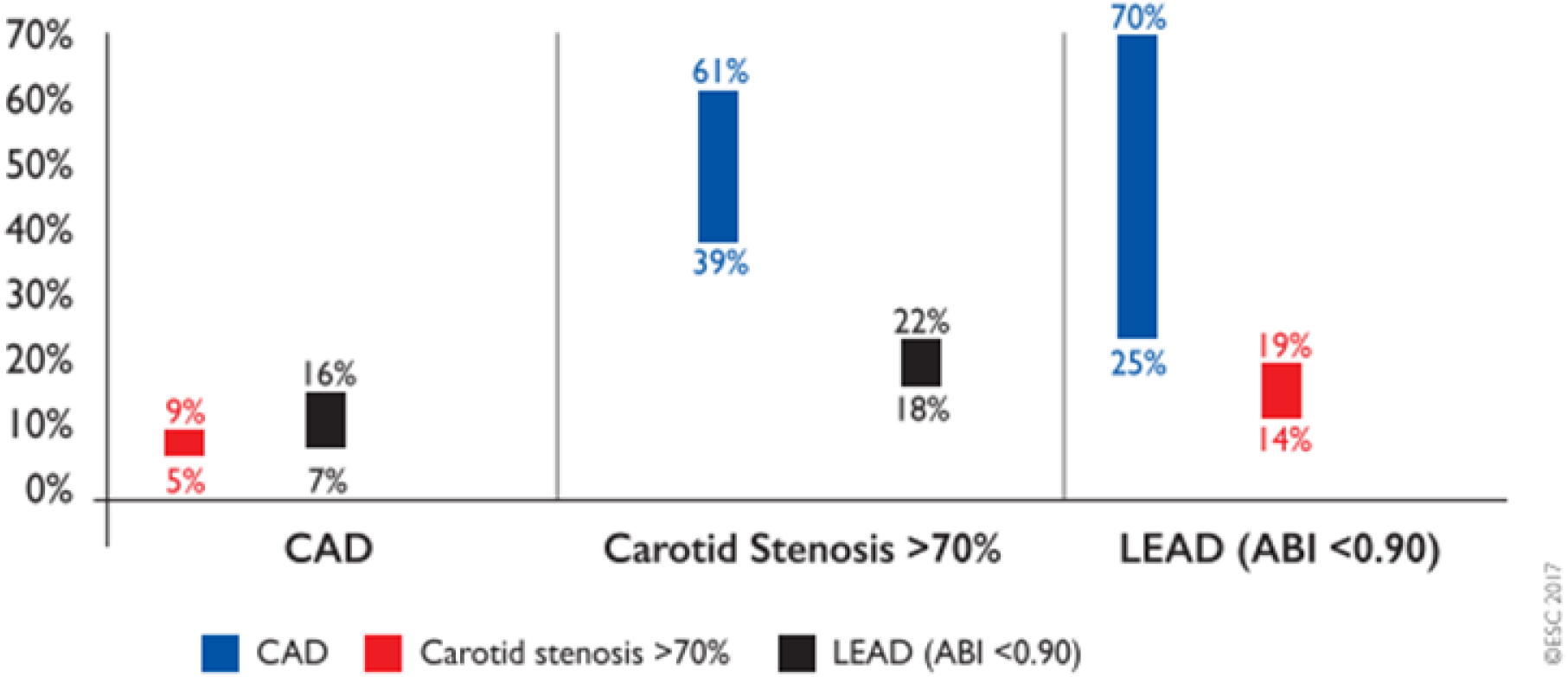
Reported rate ranges of other localizations of atherosclerosis in patients with a specific arterial disease. The graph reports the rates of concomitant arterial diseases in patients representing an arterial disease in one territory (e.g. 5-9% of patients with CAD have concomitant carotid stenosis >70%). CAD – coronary artery disease; LEAD – lower-extremity artery disease; ABI – ankle-brachial index. *Modified from ESC Clinical Practice Guidelines - Peripheral Arterial Diseases (Diagnosis and Treatment of) Guidelines* (1)

According to data from AMERICA (Aggressive detection and Management of the Extension of atherothrombosis in high Risk coronary patients In comparison with the standard of Care for coronary Atherosclerosis) 5-9% of patients with proven coronary atherosclerosis have concomitant significant carotid stenosis. 7-16% of the same population have concomitant chronic arterial disease of the limbs. 39-61% of patients with leading carotid stenosis have concomitant coronary artery disease, and 18-22% of them have concomitant arterial disease of the lower extremities. It is interesting that in patients with leading LEAD a large proportion of coronary atherosclerosis is found - 25-72%. Accompanying carotid stenosis occurs in 14-19% of this population. (1)

It is important to note the impact of atherosclerosis on the daily life of each individual. Therefore, the right approach is needed in patients with a proven outbreak of this disease and the search for “hidden” other manifestations.

## Data Availability

All data is available online.

## References

1. ESC Clinical Practice Guidelines - Peripheral Arterial Diseases (Diagnosis and Treatment of) Guidelines

2. ESC Clinical Practice Guidelines - 2019 Guidelines on Dyslipidaemias (Management of)

3. Steg PG, Bhatt DL, Wilson PW et al. One-year cardiovascular event rates in outpatients with atherothrombosis. JAMA, 2007; 297 (11): 1197–1206.

4. Fowkes FG, Low LP, Tuta S, Kozak J. Ankle-brachial index and extent of atherothrombosis in 8891 patients with or at risk of vascular disease: results of the international AGATHA study. Eur Heart J, 2006; 27: 1861–1867.

5. Hertzer NR, Beven EG, Young JR et al. Coronary artery disease in peripheral vascular patients. A classification of 1000 coronary angiograms and results of surgical management. Ann Surg, 1984; 199 (2): 223–233.

6. Eagle KA, Rihal CS, Foster ED et al. Long-term survival in patients with coronary artery disease: importance of peripheral vascular disease. The Coronary Artery Surgery Study (CASS) Investigators. J Am Coll Cardiol 1994; 23 (5):1091–1095

7. 2019 Guidelines on Chronic Coronary Syndromes ESC Clinical Practice Guidelines

8. ABC of Interventional Cardiology by Ever D. Grech.

9. Vascular imaging – I.Simova, PhD

10. I. Tasheva, I. Petrov, A. Spasov, Tsonev, S. Stankov, S. Pavlova and L. Grozdinski University Multiprofile Hospitale for Active Treatment “Acibadem City Clinic” – Sofia - CLINICAL CASES OF MULTIFOCAL ATHEROSCLEROSIS. APPROACHES FOR ENDOVASCULAR TREATMENT - Българска кардиология том XXIII, 2017, № 2

11. D. Lukanova, N. Nikolov, M. Stankev, M. Pavlov-Ultrasonographic diagnose of unstable carotid plaque -https://publishing.arbilis.com/wp-content/uploads/2015/01/NK4_3.pdf

12. prof. L.Grozdinski, m.d., MD info – 1/2015 – Multifocal aterosclerosis

13. Douglas P. Zipes MD, Peter Libby MD PhD, Robert O. Bonow MD MS, Douglas L. Mann MD, Gordon F. Tomaselli MD - Braunwald’s Heart Disease A Textbook of Cardiovascular Medicine, Single Volume

